# 24-hour Physical Activity, Sedentary, and Sleep Profiles in Individuals with Cancer: A UK Biobank Cohort Study

**DOI:** 10.1101/2025.02.25.25322841

**Authors:** Flora Le, Dorothea Dumuid, Olivia D’Elia, Alexander Haussmann, Emmanuel Stamatakis, Aiden Doherty, Yue Liao, Joshua F. Wiley

## Abstract

**Background:** The 24h behaviour profile, including physical activity, sedentary time, and sleep, is disrupted following a cancer diagnosis and contributes to cancer-related outcomes. This study describes the 24h behaviour profiles of individuals with and without cancer.

**Methods:** Seven days of accelerometer data from the UK Biobank (*M±SD_age_* = 62.3 ± 7.8y; 56.4% female) were derived by machine learning models to assess the 24h behaviour profile in individuals with cancer (n = 10 152; *M±SD_years since diagnosis_* = 7.4 ± 6.1y) compared to individuals without cancer at their accelerometry measurement (including individuals with other conditions [n = 66 403], and healthy [n = 14 726]). Diagnoses were identified through ICD-10 coding within cancer registries, hospital inpatient records, self-reports, primary care, and death registers. Bayesian compositional data analysis compared 24h behaviour profiles between individuals with and without cancer, considering time since cancer diagnosis and cancer type.

**Results:** The 24h behaviour profile of individuals with cancer consisted of 21.6 minutes in moderate-to-vigorous physical activity (MVPA), 303.5 minutes in light physical activity (LPA), 559.2 minutes in sedentary, and 555.7 minutes in sleep. Overall, they spent less time in MVPA -7.1 [-8.0, -6.1] min/day) and LPA -11.2 [-14.7, -7.7] min/day), accompanied by more sedentary time (+11.0 [7.2, 14.9] min/day) and sleep (+7.3 [4.6, 10.0] min/day), compared to healthy individuals. The least physically active profiles were observed within 1 year of cancer diagnoses and in individuals with blood, gastrointestinal tract, and lung cancer.

**Conclusions:** The 24h behaviour profiles differed by cancer history, time since diagnosis, and type. Specific behavioural trade-off strategies and support should be considered for cancer survivors, particularly in the year after diagnosis.

## Introduction

Cancer is the second leading cause of death worldwide^1^, and annual incidence rates are projected to increase by 47% for the next two decades^2^. At the same time, an aging population and longer survival times are resulting in a growing population of cancer survivors^3^. Physical inactivity is a major risk factor for comorbidities^4^, cancer recurrence^5^, and cancer mortality^5–7^. However, individuals with cancer are reported to consistently engaged in lower physical activity level^8^ than healthy individuals throughout all phases of treatment and stages of the illness^9^.

Optimising physical activity is a main target of policies and guidelines for individuals with cancer^8^. However, physical activity should not be considered as an independent behaviour. Changing time in physical activity requires trade-offs with other behaviours within the 24h day. This is because each day consists of exhaustive and mutually exclusive behaviours that always sum to 24 hours, including moderate-to-vigorous physical activity (MVPA), light physical activity (LPA), sedentary, and sleep. Among these behaviours, not only physical activity, but both longer sedentary time^10^ and short sleep (<5h)^11^ predict all-cause mortality in cancer survivors. Accordingly, the relative distribution of time across all these behaviours is more relevant than the absolute time spent in any single behaviour in isolation^12–14^. For example, an individual who reduces time in MVPA must compensate by spending more time in other behaviours (LPA, SB, and/or sleep period); they cannot decrease time in MVPA while keeping the time in other behaviours fixed. Understanding how individuals allocate their time across the full spectrum of 24h behaviours following a cancer diagnosis is crucial to the development of meaningful and targeted behaviour guidelines and interventions, that are feasible for this population and fit within the confines of a 24h day.

Analysing 24h behaviours requires compositional data analysis (CoDA)^12–14^, a methodological approach that captures the relative information of 24h behaviours and allows them to simultaneously be in one model. To our best knowledge, no studies have characterised the time allocation across 24h behaviours in individuals with cancer using CoDA. In addition, existing guidelines on 24h behaviours have relied on self-reports, which could be limited by random error and systematic bias. Evidence from objective methods, such as accelerometers, can reduce misclassification of physical activity levels and better guide personalised interventions. This study leverages the large accelerometer data from the UK Biobank (UKB) study to systematically describe the 24h behaviour profile of individuals with and without cancer. We also evaluate how the 24h behaviour profile differs according to time since cancer diagnosis and cancer types.

## Methods

### Study design and participants

The UKB is a population-based, prospective study conducted across the United Kingdom, with approximately 500 000 participants aged 40–69 years recruited in 2006–2010. We used data from a subset of 103 720 participants who provided accelerometer data in 2013–2015^15^. Raw accelerometer data from 103 626 participants were processed, excluding study withdrawals. Participants with device calibration or data reading errors (>1% of values outside +/−8g range), inadequate wear time (<72 hours), unreasonably high average acceleration (>100 mg) were excluded. Participants with any incidences between accelerometer measurement and up to one year follow-up were excluded to account for reverse causality. Those with missing healthcare linkages or covariate data, were also excluded. Participant flow diagram is in Supplementary Table S2. We followed the Strengthening the Reporting of Observational Studies in Epidemiology (STROBE)^16^ guideline (see Supplementary Figure S1).

### Cancer and non-cancer incidence ascertainment

#### Cancer group

Cancer incidence was identified using the cancer registries from England, Scotland, and Wales. We defined the *Cancer* group as those with one or more cancer incidences at the time of accelerometer measurement. *Time since cancer diagnosis* was calculated as time between the date of diagnosis and the first day of accelerometer measurement; individuals were categorised into three groups: <1 year, 1-5 years, and >5 years since cancer diagnosis. The 14 *Cancer types* commonly occurred in the UKB were also identified (see Supplementary Table S2 for ICD-10 codes), including *Blood, Breast, Colorectal, Endocrine Gland, Gastrointestinal, Genitourinary, Gynaecological, Head and Neck, Lung, Melanoma, Skin (non-melanoma), Prostate, Multiple Primary (more than one type), and Other Cancer*. Participants with cancer who have one or more other conditions were included in the cancer group, due to high prevalence of multimorbidity with cancer (94% in the current study, consistent with 91% in a previous study^17^).

#### Non-cancer group

Non-cancer incidence was identified using hospital inpatient records, self-reports, primary care, and death registers. The incidences were collated into 11 chronic conditions defined according to International Classification of Diseases (ICD-10) Chapters (see Supplementary Table S3 for ICD-10 codes). We defined two Non-cancer groups, 1) *Healthy* as no incidence, and 2) *Other Conditions* as one or more chronic conditions (except cancer) at the time of accelerometry.

#### 24h behaviour assessment

MVPA, LPA, sedentary, and sleep period were derived from the Axivity AX3 wrist-worn triaxial accelerometers. Participants were instructed to wear the device on their dominant wrist continuously for seven days. The accelerometers captured three-dimensional acceleration data at 100Hz with a dynamic range of ± 8g. Accelerometer data were extracted and processed using methods described previously^15^. Time spent in 24h behaviours were calculated using random forest and hidden Markov model machine-learning methods^18^.

### Statistical analysis

We used a Bayesian compositional analysis approach. The raw 4-part composition of the 24h behaviours (MVPA, LPA, sedentary behaviour, and sleep period), were expressed as a set of three *isometric log ratio* (*ilr)* coordinates for model estimation. This *ilr* transformation, using a sequential binary partition^19^, prevents the perfect multicollinearity associated with fitting raw compositional data.

To determine the association between *Time since cancer diagnosis* and 24h behaviour profile, a Bayesian multivariate model (a) was fitted with the *ilr* coordinates representing the 24h behaviour profile as the outcome and *Time since cancer diagnosis* (5 categories consisting of 3 cancer groups [<1 year, 1-5 years, >5 years], 2 non-cancer groups [*Other Conditions* and *Healthy*]) as the predictors. Time in 24h behaviours in the *Cancer* group was calculated as the weighted mean of the three cancer groups. Post-hoc contrasts were used to estimate the differences in 24h behaviours by *Time since cancer diagnosis* compared to the two non-cancer groups. Two methods of contrasts were employed 1) treatment versus control, where *Time since cancer diagnosis* was compared against the *Healthy* and *Other Conditions* groups, and 2) pairwise comparisons within *Cancer* group, a) <1 year versus 1-5 years; b) <1 years versus >5 years; and c) 1-5 years versus >5 years.

To assess *Cancer types* and 24h behaviour profile, a Bayesian multivariate model (b) was fitted with the *ilr* coordinates representing the 24h behaviour profile as the outcome and *Cancer types* (16 categories of 14 cancer types, and 2 non-cancer groups) as the predictors. Post-hoc contrasts, using the treatment versus control method, estimated the difference in the time spent in 24h behaviours between each of the 14 cancer types and the two control groups (*Healthy* and *Other Conditions*), respectively.

In line with previous studies^20–22^, model (a) adjusted for age at accelerometry, sex, ethnicity, Townsend deprivation index, education/qualification attainment, employment, smoking status, and alcohol consumption (see Supplementary Table S1 for directed acyclic graph). Model (b) adjusted for the same covariates plus *time since cancer diagnosis* as this may vary by cancer type. All continuous covariates were modelled using smooth functions to account for possible non-linear associations. Both models were fitted with weakly-informative priors and 4 chains with 2000 iterations including (500 warmups) per chain for a total of 6000 post-warmup draws. Model convergence was confirmed (all 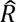 < 1.05 and effective sample size > 400)^23^. Bayesian posterior means and 99% credible interval were reported. Analyses were performed in R using package multilevelcoda^24,25^ and brms^26^.

## Results

The analytical sample included 91 281 individuals [average age at accelerometer measurement (SD) = 62.30 (7.85) years; 56.4% female], details in Table 1. There were 10 152 (11.1%) participants in the *Cancer* group, 66 403 (72.7%) in the *Other Conditions* group, and 14 726 (16.1%) in the *Healthy* group. Individuals with *Cancer* provided accelerometer data after 7.4 (SD = 6.1) years since cancer diagnosis. They had an average of 2.56 (1.80) other major chronic conditions by categories defined in the ICD-10, comparable with the *Other Conditions* group with 2.58 (1.53) conditions. Quantiles of 24h behaviour profile in individuals with cancer by demographic subgroups are reported in Supplementary Table S5.

**Table 1.** Participant characteristics (*N* = 91 352).

| | **Cancer**<br>($n = 10\,152$) | **Other Conditions**<br>($n = 67\,478$) | **Healthy**<br>($n = 13\,722$) |
| --- | --- | --- | --- |
| **AT RECRUITMENT** |  |  |  |
| **Age $M$ ($SD$)** | 59.66 (6.79) | 56.35 (7.72) | 52.63 (7.64) |
| **Sex $N$ (%)** |  |  |  |
| Female | 5 660 (55.8%) | 37 955 (56.2%) | 7 904 (57.6%) |
| Male | 4 492 (44.2%) | 29 523 (43.8%) | 5 818 (42.4%) |
| **White ethnicity $N$ (%)** | 10 016 (98.7%) | 65 412 (96.9%) | 13 215 (96.3%) |
| **Body mass index $N$ (%)** |  |  |  |
| Under to Normal weight ($< 25\text{ kg/m}^2$) | 3942 (38.9%) | 24935 (37.0%) | 6973 (50.9%) |
| Overweight ($25.0$ to $29.9\text{ kg/m}^2$) | 4251 (42.0%) | 28131 (41.8%) | 5187 (37.8%) |
| Obese ($30+$ $\text{kg/m}^2$) | 1940 (19.1%) | 14255 (21.2%) | 1547 (11.3%) |
| **Education/qualification attainment $N$ (%)** | 9 073 (89.4%) | 61 572 (91.2%) | 13 125 (95.6%) |
| **Employment $N$ (%)** | 5 563 (54.8%) | 45 928 (68.1%) | 11 331 (82.6%) |
| **Townsend deprivation index $N$ (%)** |  |  |  |
| Least deprived ($< -3.8$) | 2 755 (27.1%) | 16 959 (25.1%) | 3 423 (24.9%) |
| Second least deprived ($-3.8$ to $-2.5$) | 2 557 (25.2%) | 16 010 (23.7%) | 3 239 (23.6%) |
| Second most deprived ($-2.5$ to $-0.2$) | 2 640 (26.0%) | 17 462 (25.9%) | 3 457 (25.2%) |
| Most deprived ($> -0.2$) | 2 200 (21.7%) | 17 047 (25.3%) | 3 603 (26.3%) |
| **Smoking $N$ (%)** |  |  |  |
| Never | 45 034 (67.8%) | 38 025 (56.4%) | 8 676 (63.2%) |
| Previous | 4 030 (39.7%) | 24 794 (36.7%) | 4 059 (29.6%) |
| Current | 606 (6.0%) | 46 59 (6.9%) | 987 (7.2%) |
| **Alcohol consumption $N$ (%)** |  |  |  |
| Never | 273 (2.7%) | 2 014 (3.0%) | 361 (2.6%) |
| Previous | 259 (2.6%) | 2 006 (3.0%) | 227 (1.7%) |
| Current | 9 620 (94.8%) | 63 458 (94.0%) | 13 134 (95.7%) |
| **AT THE START OF ACCELEROMETRY** |  |  |  |
| **Age $M$ ($SD$)** | 65.89 (6.79) | 62.49 (7.74) | 58.72 (7.69) |
| **Other Non-cancer Conditions (by ICD chapters)** |  |  | — |
| **$N$ (%)** | 9 456 (93.1%) |  |  |
| **$M$ ($SD$)** | 2.56 (1.80) | 2.58 (1.53) |  |
| **Time since cancer diagnosis $N$ (%)** |  | — | — |
| $< 1$ year | 971 (9.6%) | | |
| $1$ to $5$ years | 3 451 (34.0%) | | |
| $> 5$ years | 5 730 (56.4%) | | |
| **Cancer type $N$ (%), median time since diagnosis** |  | — | — |
| Blood | 412 (4.1%), 7.40 |  |  |
| Breast | 1 911 (18.8%), 9.23 |  |  |
| Colorectal | 391 (3.9%), 6.54 |  |  |
| Endocrine Gland | 79 (0.8%), 9.47 |  |  |
| Gastrointestinal Tract | 100 (1.0%), 5.93 |  |  |
| Genitourinary | 296 (2.9%), 9.95 |  |  |
| Gynaecological | 401 (3.9%), 9.25 |  |  |
| Head and Neck | 115 (1.1%), 7.53 |  |  |
| Lung | 57 (0.6%), 5.46 |  |  |
| Melanoma | 395 (3.9%), 9.08 |  |  |
| Multiple Primary | 1 714 (16.9%), 6.44 |  |  |
| Other Cancer | 148 (1.5%), 10.73 |  |  |
| Skin (non-melanoma) | 3 017 (29.7%), 6.85 |  |  |
| Prostate | 1 116 (11.0%), 5.10 |  |  |
| **Minutes in 24h behaviours $M$ ($SD$)** |  |  |  |
| Moderate-to-vigorous physical activity | 38.3 (32.8) | 40.2 (34.3) | 48.5 (36.5) |
| Light physical activity | 296.7 (95.8) | 302.8 (98.8) | 308.3 (98.2) |
| Sedentary behaviour | 566.4 (107.6) | 565.6 (109.6) | 557.0 (107.2) |
| Sleep period | 538.7 (77.0) | 531.5 (74.8) | 526.2 (69.8) |

Notes. 24h behaviours are presented as arithmetic means. Percentages may not sum to 100% due to rounding.

### 24h profiles of individuals with and without cancer

Across the three groups, participants allocated most of their day in sedentary time, followed by sleep period, LPA, and lastly MVPA (Figure 1). The 24h behaviour profile of individuals with *Cancer* consisted of 559.2 [99%CI 553.8, 564.4] minutes in sedentary behaviour, 555.7 [552.0, 559.5] minutes in sleep period, 303.5 [298.8, 308.5] minutes in LPA and 21.6 [20.6, 22.6] minutes in MVPA. Individuals with *Other Conditions* allocated 557.5 [553.0, 561.7] minutes in sedentary behaviour, 550.7 [547.6, 553.9] minutes in sleep period, 309.5 [305.5, 313.7] minutes in LPA and 22.3 [21.5, 23.2] minutes in MVPA, whereas *Healthy* individuals allocated 548.1 [543.1, 553.0] minutes in sedentary behaviour, 548.4 [545.1, 551.9] minutes in sleep period, 314.8 [310.2, 319.5] minutes in LPA and 28.6 [27.4, 29.9] minutes in MVPA. The estimated 24h behaviour profile of individuals with *Cancer* showed more similarity to that of individuals with *Other Conditions* than to that of individuals who were classified as *Healthy*.

**Figure 1.**
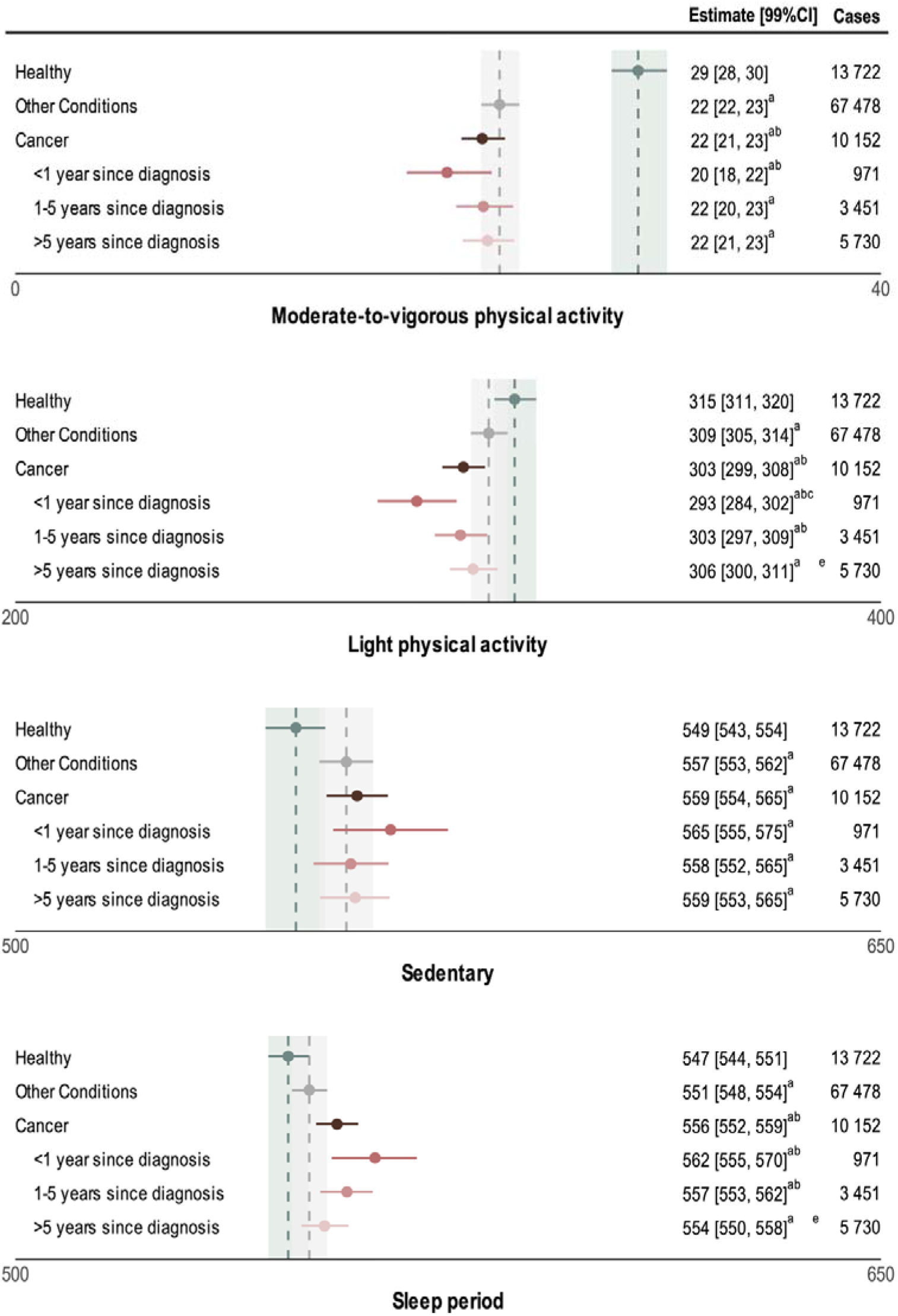
Estimated 24h behaviour profile (in minutes) across time since cancer diagnosis. Values are Bayesian posterior mean differences and 99% credible intervals. Model adjusted for age at accelerometer, sex, ethnicity, Townsend deprivation index, education/qualification attainment, employment, smoking status, and alcohol consumption. Vertical dashed lines represent estimated means, and shaded regions are 99% credible intervals, of non-cancer groups (green = healthy, grey = other conditions). ^a^significant difference from *Healthy*, ^b^significant difference from *Other Conditions*, ^c^significant difference between <1 year and 1-5 years since diagnosis, ^d^significant difference between 1-5 and >5 years since diagnosis, ^e^significant difference between <1 year and >5 years since diagnosis.

### Differences in 24h behaviour profiles across time since cancer diagnosis

Throughout time since cancer diagnoses, individuals with *Cancer* showed significant differences in their 24h behaviours compared to *Healthy* individuals (Table 2 and Figure 1). Specifically, individuals with *Cancer* spent less time in MVPA (-7.06 [99%CI -8.02, -6.09] minutes) and LPA (-11.24 [-14.74, -7.73] minutes) and more in sedentary time (+11.02 [7.19, 14.89] minutes) and sleep period (+7.28 [4.62, 9.95] minutes). The largest differences in 24h behaviour profiles were observed in individuals <1 year since cancer diagnosis, followed by 1-5 years since cancer diagnosis. Individuals who were >5 year since cancer diagnosis had a 24h profile more similar to that of *Healthy* individuals.

**Table 2.** Estimated minute differences in 24h behaviour profile across time since cancer diagnosis. Values are Bayesian posterior mean differences and 99% credible intervals. Model adjusted for age at accelerometer, sex, ethnicity, Townsend deprivation index, education/qualification attainment, employment, smoking status, alcohol consumption, and other chronic conditions. MVPA = moderate-to-vigorous physical activity, LPA = light physical activity. Bolded values indicate 99% credible intervals not containing 0.

|  | MVPA | LPA | Sedentary | Sleep Period |
| --- | --- | --- | --- | --- |
| **ref. Healthy** |  |  |  |  |
| **Cancer** | **-7.21**<br>[-8.21, -6.27] | **-11.85**<br>[-15.45, -8.36] | **10.60**<br>[6.76, 14.47] | **8.46**<br>[5.67, 11.18] |
| <1 year since diagnosis | **-8.84**<br>[-10.76, -6.88] | **-22.62**<br>[-31.09, -14.05] | **16.41**<br>[7.1, 25.67] | **15.05**<br>[7.95, 21.87] |
| 1-5 years since diagnosis | **-7.16**<br>[-8.4, -5.84] | **-12.53**<br>[-17.68, -7.36] | **9.51**<br>[3.8, 15.22] | **10.18**<br>[6.36, 14.07] |
| >5 year since diagnosis | **-6.97**<br>[-8.13, -5.86] | **-9.61**<br>[-13.9, -5.45] | **10.28**<br>[5.76, 15.02] | **6.30**<br>[3.12, 9.51] |
| **ref. Other Conditions** |  |  |  |  |
| **Cancer** | **-0.80**<br>[-1.47, -0.13] | **-5.86**<br>[-8.75, -3.02] | 1.86<br>[-1.21, 4.93] | **4.80**<br>[2.57, 6.90] |
| <1 year since diagnosis | **-2.43**<br>[-4.15, -0.57] | **-16.63**<br>[-24.82, -8.14] | 7.66<br>[-1.37, 16.58] | **11.39**<br>[4.57, 18.2] |
| 1-5 years since diagnosis | -0.75<br>[-1.79, 0.32] | **-6.54**<br>[-11.09, -1.60] | 0.77<br>[-4.18, 5.99] | **6.52**<br>[3.03, 10.06] |
| >5 year since diagnosis | -0.56<br>[-1.44, 0.36] | -3.62<br>[-7.30, 0.06] | 1.53<br>[-2.25, 5.54] | 2.65<br>[-0.19, 5.45] |
| **Pairwise across time since diagnosis** |  |  |  |  |
| 1-5 years since diagnosis<br>ref. <1 year since diagnosis | 1.68<br>[-0.51, 3.81] | **10.09**<br>[0.59, 19.36] | -6.90<br>[-17.26, 3.55] | -4.87<br>[-12.34, 2.79] |
| >5 year since diagnosis<br>ref. 1-5 years since diagnosis | 0.19<br>[-1.15, 1.5] | 2.92<br>[-2.85, 8.5] | 0.77<br>[-5.26, 6.83] | -3.87<br>[-8.09, 0.36] |
| >5 year since diagnosis<br>ref. <1 year since diagnosis | 1.87<br>[-0.16, 3.79] | **13.00**<br>[3.77, 22.02] | -6.13<br>[-16.1, 3.6] | **-8.74**<br>[-16.11, -1.53] |

For all *Time since cancer diagnosis* categories, individuals with *Cancer* allocated their time differently among MVPA, LPA, and sleep period, compared to individuals with *Other Conditions*, whereas their sedentary time was similar (Table 2 and Figure 1). Specifically, individuals with *Cancer* spent on average, less time in MVPA (-0.76 [-1.42, -0.09] minutes) and LPA (-5.93 [-8.76, -3.1] minutes), and more time in sleep period (+5.01 [2.82, 7.14] minutes). The largest differences in 24h behaviour profiles were observed in individuals <1 year since cancer diagnosis, then 1-5 years since cancer diagnosis. Individuals who were >5 year since cancer diagnosis showed a similar composition to that of individuals with *Other Conditions,* with the only difference found for sleep period (+2.86 [0.08, 5.59] minutes).

### Differences in 24h behaviours across cancer types

Individuals with different *Cancer types* had different 24h composition compared to *Non-cancer* individuals, including those who are *Healthy* and those with *Other Conditions* (Table 3 and Figure 2). Compared to *Healthy* individuals, all *Cancer types* were consistently associated with less time in MVPA, and several *Cancer types* were additionally associated with less time in LPA and more sedentary time. Compared to individuals with *Other Conditions*, several *Cancer types* were associated with less time in LPA, followed by MVPA, and more sedentary time. Only some *Cancer types* had different sleep periods compared to *Non-cancer* groups.

**Figure 2.**
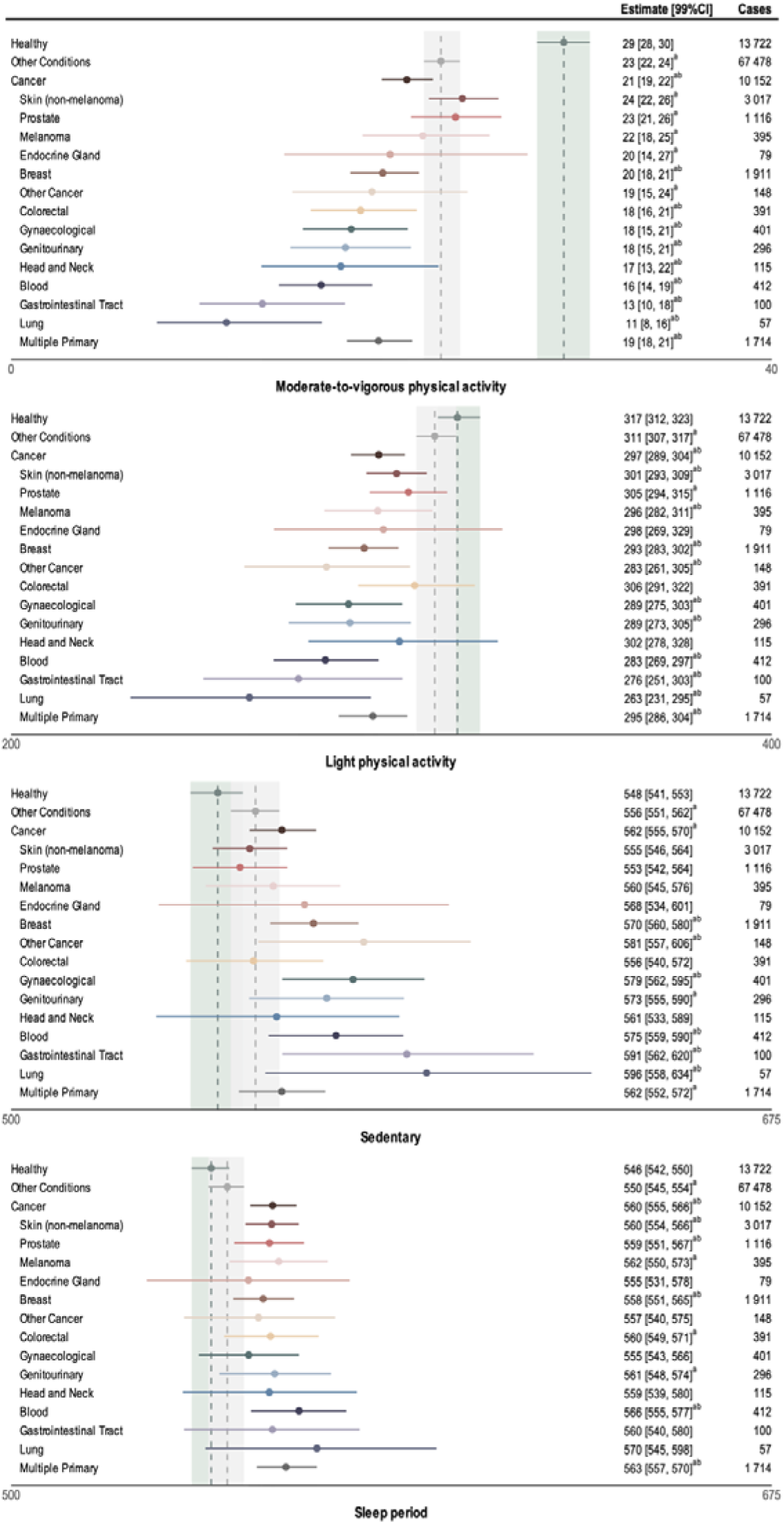
Estimated 24h behaviour profile (in minutes) across cancer types. Values are Bayesian posterior mean differences and 99% credible intervals. Model adjusted for age at accelerometer, sex, ethnicity, Townsend deprivation index, education/qualification attainment, employment, smoking status, alcohol consumption, and time since cancer diagnosis. Vertical dashed lines represent estimated means, and shaded regions are 99% credible intervals, of non-cancer groups (green = healthy, grey = other conditions). ^a^significant difference from *Healthy*, ^b^significant difference from *Other Conditions*.

**Table 3.** Estimated minute differences in 24h behaviour profile in across cancer types. Values are Bayesian posterior mean differences and 99% credible intervals. Model adjusted for age at accelerometer, sex, ethnicity, Townsend deprivation index, education/qualification attainment, employment, smoking status, and alcohol consumption. MVPA = moderate-to-vigorous physical activity, LPA = light physical activity. Bolded values indicate 99% credible intervals not containing 0.

|  | MVPA | LPA | Sedentary | Sleep Period |
| --- | --- | --- | --- | --- |
| **ref. Healthy** |  |  |  |  |
| Blood | **-12.76**<br>[-15.33, -9.87] | **-34.68**<br>[-49.76, -20.35] | **27.23**<br>[11.33, 44.13] | **20.22**<br>[9.02, 31.86] |
| Breast | **-9.52**<br>[-11.58, -7.47] | **-24.48**<br>[-35.75, -14.65] | **22.04**<br>[11.17, 34.01] | **11.96**<br>[4.05, 20.31] |
| Colorectal | **-10.68**<br>[-13.54, -7.56] | -11.20<br>[-26.61, 4.92] | 8.23<br>[-7.87, 25.50] | **13.65**<br>[2.06, 25.59] |
| Endocrine Gland | **-9.13**<br>[-14.78, -2.00] | -19.42<br>[-48.84, 12.06] | 19.99<br>[-13.23, 53.23] | 8.56<br>[-14.92, 32.17] |
| Gastrointestinal Tract | **-15.85**<br>[-19.48, -11.42] | **-41.76**<br>[-66.77, -14.92] | **43.53**<br>[14.48, 72.49] | 14.09<br>[-6.71, 34.50] |
| Genitourinary | **-11.48**<br>[-14.76, -7.94] | **-28.22**<br>[-45.26, -11.84] | **25.09**<br>[6.52, 43.65] | 14.61<br>[1.48, 28.49] |
| Gynaecological | **-11.18**<br>[-14.09, -8.18] | **-28.59**<br>[-43.52, -13.81] | **31.15**<br>[14.5, 48.26] | 8.62<br>[-3.2, 20.47] |
| Head and Neck | **-11.72**<br>[-16.2, -6.49] | -15.21<br>[-39.83, 10.36] | 13.56<br>[-14.65, 42.34] | 13.37<br>[-6.57, 33.43] |
| Lung | **-17.74**<br>[-21.68, -12.61] | **-54.67**<br>[-86.5, -21.31] | **48.08**<br>[10.63, 85.66] | 24.34<br>[-1.62, 52.36] |
| Melanoma | **-7.41**<br>[-10.71, -3.66] | **-20.91**<br>[-36.11, -5.76] | 12.77<br>[-3.46, 29.42] | **15.55**<br>[3.47, 27.88] |
| Multiple Primary | **-9.74**<br>[-11.74, -7.87] | **-22.19**<br>[-32.66, -12.67] | **14.73**<br>[4.37, 26.14] | **17.20**<br>[9.66, 25.29] |
| Other Cancer | **-10.09**<br>[-14.33, -4.93] | **-34.40**<br>[-56.93, -12.17] | **33.61**<br>[8.57, 59.47] | 10.87<br>[-6.59, 28.94] |
| Skin (non-melanoma) | **-5.34**<br>[-7.36, -3.42] | **-15.94**<br>[-26.12, -7.48] | 7.36<br>[-1.85, 18.28] | **13.92**<br>[7.19, 21.46] |
| Prostate | **-5.70**<br>[-8.21, -3.10] | **-12.83**<br>[-24.67, -2.05] | 5.12<br>[-6.45, 17.64] | **13.41**<br>[5.01, 22.84] |
| **ref. Other Conditions** |  |  |  |  |
| Blood | **-6.29**<br>[-8.62, -3.64] | **-28.75**<br>[-43.67, -14.79] | **18.54**<br>[2.66, 35.3] | **16.51**<br>[5.58, 28.48] |
| Breast | **-3.05**<br>[-4.91, -1.22] | **-18.55**<br>[-29.57, -8.92] | **13.34**<br>[3.08, 25.62] | **8.26**<br>[0.67, 16.48] |
| Colorectal | **-4.22**<br>[-6.95, -1.31] | -5.26<br>[-20.71, 10.46] | -0.46<br>[-16.23, 16.80] | 9.94<br>[-1.37, 21.66] |
| Endocrine Gland | -2.66<br>[-8.16, 4.52] | -13.49<br>[-42.79, 17.92] | 11.29<br>[-22.01, 44.61] | 4.86<br>[-18.64, 28.64] |
| Gastrointestinal Tract | **-9.39**<br>[-12.91, -5.08] | **-35.83**<br>[-60.51, -8.74] | **34.84**<br>[5.63, 63.85] | 10.38<br>[-10.17, 30.35] |
| Genitourinary | **-5.01**<br>[-8.06, -1.45] | **-22.29**<br>[-39.57, -5.73] | 16.40<br>[-2.07, 34.81] | 10.90<br>[-2.31, 24.52] |
| Gynaecological | **-4.71**<br>[-7.43, -1.77] | **-22.66**<br>[-37.77, -8.03] | **22.46**<br>[5.72, 39.41] | 4.92<br>[-6.92, 16.42] |
| Head and Neck | **-5.26**<br>[-9.59, -0.06] | -9.27<br>[-33.91, 16.42] | 4.86<br>[-23.5, 32.96] | 9.67<br>[-10.23, 29.90] |
| Lung | **-11.28**<br>[-15.01, -6.23] | **-48.74**<br>[-80.06, -15.31] | **39.39**<br>[1.93, 77.65] | 20.63<br>[-5.40, 48.55] |
| Melanoma | -0.94<br>[-4.20, 2.71] | **-14.98**<br>[-30.15, -0.22] | 4.08<br>[-12.45, 20.73] | 11.84<br>[-0.29, 24.26] |
| Multiple Primary | **-3.27**<br>[-5.10, -1.53] | **-16.26**<br>[-26.66, -6.81] | 6.04<br>[-4.32, 17.58] | **13.50**<br>[6.09, 21.55] |
| Other Cancer | -3.62<br>[-7.82, 1.42] | **-28.47**<br>[-50.86, -6.23] | **24.92**<br>[0.21, 50.81] | 7.17<br>[-10.06, 25.35] |
| Skin (non-melanoma) | 1.13<br>[-0.63, 2.96] | **-10.00**<br>[-19.85, -1.64] | -1.34<br>[-10.25, 9.05] | **10.21**<br>[3.61, 17.68] |
| Prostate | 0.77<br>[-1.65, 3.20] | -6.90<br>[-18.51, 3.57] | -3.57<br>[-14.89, 8.83] | **9.71**<br>[1.46, 18.76] |

Among *Cancer types*, individuals with *Blood, Gastrointestinal Tract,* and *Lung Cancer* showed the largest difference in their 24h behaviour profiles compared to *Non-cancer* individuals (Table 3 and Figure 2). Compared to *Healthy* individuals, individuals with these three *Cancer types* spent less time in MPVA (estimated minute differences range between -12.58 [-15.11, -9.73] and -17.54 [-21.59, -12.54]) and LPA (estimated minute differences range between -33.99 [-48.66, -19.06] and -53.92 [-85.22, -20.57]). They also had more sedentary time (estimated minute differences range between +27.55 [11.62, 43.61] and +48.22 [11.03, 85.50]) and longer sleep period (estimated minute differences range between +19.01 [7.15, 30.88] and +23.24 [-3.49, 51.57]; statistically significant for *Blood,* not *Gastrointestinal Tract,* and *Lung Cancer*). Similar patterns emerged in comparison to individuals with *Other Conditions,* but the magnitudes were smaller.

Among *Cancer types*, individuals with *Skin (non-melanoma), Prostate,* and *Melanoma Cancer* showed the smallest difference in their 24h composition from *Non-cancer* individuals (Table 3 and Figure 2). Despite having the smallest difference, these individuals had less time in MPVA (estimated minute differences range between -5.17 [-7.37, -3.19] and -7.25 [-10.67, -3.48]) and LPA (estimated minute differences range between -12.29 [-23.54, -1.94] and -20.37 [-36.45, -4.76]), and a longer sleep period (estimated minute differences range between +12.23 [3.63, 21.26] and +14.51 [2.45, 26.89]). Sedentary time was not different (estimated minute differences range between +5.63 [-5.83, 17.80] and +13.12 [-3.41, 29.83]). Similar patterns emerged in comparison to individuals with *Other Conditions*, but the estimates were smaller.

## Discussion

In this large cohort study, we observed evidence of 24h behaviour disruption in individuals with *Cancer*. Overall, individuals with *Cancer* spent less time in MVPA and LPA, and compensated by more sedentary time and longer sleep periods, compared to *Non-cancer* individuals, including those are *Healthy* and those with *Other Conditions*. We also identified that the largest disruptions in 24h behaviours were observed in those within 1 year since diagnosis. For almost every *Cancer type,* the 24h behaviour profile deviated from that observed in *Non-cancer*, with individuals with *Blood, Gastrointestinal Tract,* and *Lung Cancer* showing the largest differences in their 24h behaviours compared to individuals without cancer.

### 24h behaviour profile in individuals with cancer vs non-cancer

Findings highlight an overall profile of reduced physical activity regardless of intensity levels, and increased sedentary time, in individuals with *Cancer* compared to *Non-cancer*. Specifically, they had less MVPA (7 minutes less) and LPA (11 minutes less), and more sedentary time (+11 minutes) and sleep period (+7 minutes).

On average, people with cancer had 22 minutes of MVPA per day. The adverse effects of the cancer disease and treatments, including fatigue, pain, and nausea, may limit the ability of individuals to achieve current recommendations^8^ for cancer survivors, which focus on achieving the same levels of MVPA recommended for healthy individuals^27^. Our findings show that all activity behaviours are affected when people have a cancer diagnosis and suggest that a holistic approach which considers time use across the whole day is warranted. Such an approach may minimise sitting time in favour of physical activity across intensity levels for individuals with cancer, aligning with existing evidence on the reduced risk of cancer associated with engaging in vigorous intensity physical activity^22^, or MVPA and LPA at the expense of sedentary time^21^. For example, breaking up sedentary time with short bouts of physical activity at any intensity levels, could be a more feasible approach for individuals who are unable or reluctant to initiate or maintain adequate levels of MVPA.

Individuals with *Cancer* also spent more time in sleep period than *Non-cancer* individuals. The average sleep period of individuals with cancer was 9.3 hours, exceeding the 7-9 hour threshold of optimal sleep recommended for healthy adults^28^. Sleep is important for improving quality of life^29^ and mortality^30^ of individuals with cancer. However, excessive sleep may be a consequence of cancer-related fatigue, and psychological distress (depression and anxiety), and risk factor for mortality^31,32^ and incidence of other chronic conditions^32^. As sleep disturbances have been consistently reported throughout all phases of treatment and stages of cancer^33–35^, it is possible that our participants had a long sleep period but disrupted, unrestful sleep (e.g., longer sleep onset latency or night-time awakenings). Our sleep period variable only captured the time between sleep onset and offset, and did not consider night awakenings. To improve the understanding of sleep in cancer, further research should consider the multidimensional nature of sleep, including objective sleep duration and time awake in bed, and subjective sleep complaints. There is urgent need to optimally support sleep after cancer^36^ as sleep difficulties persist well after diagnosis and completion of treatment^37,38^.

### 24h behaviour profile across time since cancer diagnosis

Across the time since cancer diagnosis, the greatest deviation from the *Healthy* profile was observed in those within 1 year of cancer diagnosis. Individuals in this category engaged in 31 min less physical activity across intensity levels (9 minutes less in MVPA and 22 minutes less in LPA), and instead allocated the time to inactive/resting behaviours (17 minutes more sedentary time and 14 minutes more in sleep period), compared to *Healthy* individuals. Timely care for individuals recently diagnosed with cancer to meet physical activity guidelines might be a valuable approach to mitigate cancer-related physical and psychological symptoms^39–41^.

Individuals 1-5 years since cancer diagnosis were more physically active than those within 1 year since diagnosis, and individuals 5 years or more after cancer diagnosis showed additional improvement. The gradual improvement in 24h behaviours along the trajectory of survivorship may be due to lifestyle modifications and cancer remission. Nevertheless, individuals in the 5 years since cancer diagnosis category were not as physically active as *Non-cancer* individuals, and their 24h behaviour profile still significantly deviated from the *Healthy* profile. This trajectory of the 24h behaviours underscores the value of cancer care aimed at 24h behaviours spanning from diagnosis to long-term survivorship to meet the needs of the growing population of cancer survivors^3^.

### 24h behaviour profile across cancer types

Across cancer types, physical activity regardless of intensity levels was lower than in the *Non-cancer* group. Of the 14 cancer types examined, individuals with cancers that typically have a poorer prognosis (e.g., *Blood, Gastrointestinal Tract,* and *Lung*), were the least active, whereas those with cancers that have better prognoses, including *Skin (non-melanoma), Prostate,* and *Melanoma*, were more active. For example, between 2013 and 2019, the 5-year relative survival rate for *Lung* cancer was 25%, compared to 97% for *Prostate* cancer^32^. These findings also suggest that specific 24h behaviour profiles across cancer types may be due to the severity of the cancer disease and invasiveness of treatment. Further research incorporating information on cancer stage and treatment could provide more systematic evidence.

It is important to recognise that what is regarded as healthy might differ among individuals with cancer, especially those are diagnosed with invasive cancer and undergo aggressive treatments. For individuals with *Blood, Gastrointestinal Tract,* and *Lung* cancer, trying to achieve a physical activity guideline threshold may be difficult and have limited benefits on health outcomes and survival. A more feasible approach could be reducing sedentary time or optimising sleep. For example, psychological factors, such as depressive symptoms, intrusive and avoidant thoughts about cancer, have been found to predict long-term trajectory of sleep in cancer patients^37^. Future studies that promote the optimal dose of 24h behaviours, while addressing modifiable factors, could provide precise recommendations for specific cancer types.

### Strengths and limitations

The strengths of this study include the use of Bayesian compositional data analysis to account for the compositional nature of 24h behaviours and obtain direct uncertainty estimates, while accommodating potential non-linear associations and adjustment for several plausible confounders. The large sample size enables comparison across the progression of cancer disease, and several common and underrepresented cancer types. The wearables-based measures of behaviours overcome inherent limitations of self-report and provide useful evidence to enhance the current public guidelines on 24h behaviours. The use of wearables to track 24h behaviour trajectories holds promise to facilitate better awareness and management of cancer-related symptoms.

Despite the temporal sequence of the prospective design, the observational design of the UKB limits our analysis from probing causality or the within-person changes in 24 behaviour time allocation from pre-diagnosis to post-diagnosis. This study is also limited by the lack of information on stage and grade of the tumour, and treatment, from the UKB. Further, we did not exclude those with other conditions from the *Cancer* group, given the high prevalence of multimorbidity (94%) in the *Cancer* group. Given the cancer’s clinical complexity, it is difficult to precisely disentangle whether the other chronic conditions are contributors or consequences of cancer, and how they uniquely contribute to the 24h behaviour profile. The potential misclassification of posture and movement, which is inherent to wrist-worn devices, limits our ability to separate the effects of sleep duration versus time awake in bed, which is important for the understanding of sleep and informing care. Lastly, survival bias may have influenced the UKB data and our observations. Further studies in nationally representative cohorts should evaluate the generalisability.

### Conclusion

This study provided insights into 24h behaviour profiles among individuals with cancer, across time since cancer diagnosis and cancer types, and how they differ to profiles observed among individuals without cancer. Our study can improve progress toward optimal survivorship care delivery for the large and growing population of individuals living with and beyond cancer. Public guidelines for healthy individuals have transitioned from providing guidance for independent behaviours and towards integrating 24h behaviours^42^, but only general MVPA recommendations are available for cancer survivors^8^. Our findings are supportive of an approach towards healthy daily balance of physical activity, sedentary, and sleep, rather than one-size-fits-all target of increasing physical activity^43^ for individuals with cancer. Recommendations on 24h behaviours should be implemented ideally following a cancer diagnosis and follow-up into long-term survivorship, and tailored to cancer types. People with cancers that have poor prognoses, including *Blood*, *Lung*, and *Gastrointestinal* cancers, had the least healthy 24h behaviour profile, highlighting the need for further support and resources to be allocated to optimise survivorship care and health outcomes for these groups.

## Ethics approval

The UK Biobank has generic ethical approval from the Northwest Multi-Centre Research Ethics Committee (11/NW/03820).

## Data availability

Participants who accepted the invitation to join the UK Biobank cohort provided written, informed consent and were given reimbursement for travel expenses. Data are available on application to the UK Biobank (www.ukbiobank.ac.uk/). This research is approved for access under application #62254.

## Code availability

Analysis code for this study is available at https://github.com/florale/ukb-cancer-24h

## Acknowledgement

We thank the participants in the UK Biobank and the UK Biobank for making the data and resources available. FL was supported by Monash University through a Monash Graduate Scholarship and a Monash International Tuition Scholarship. DD was funded by an Australian Research Council (ARC) Discovery Early Career Award (DECRA) DE230101174 and by the Centre of Research Excellence in Driving Global Investment in Adolescent Health funded by National Health and Medical Research Council (NHMRC) GNT1171981. AH was funded by the Deutsche Forschungsgemeinschaft (DFG, German Research Foundation [540707655]). JFW was supported by a NHMRC fellowship (1178487). is supported by the Wellcome Trust [223100/Z/21/Z, 227093/Z/23/Z], Novo Nordisk, Swiss Re, and the British Heart Foundation Centre of Research Excellence [RE/18/3/34214]. YL is supported by the American Institute for Cancer Research [IIG 849385] and the American Heart Association [23CDA1050513]

## Supplementary Materials

**Supplementary Table S1.**
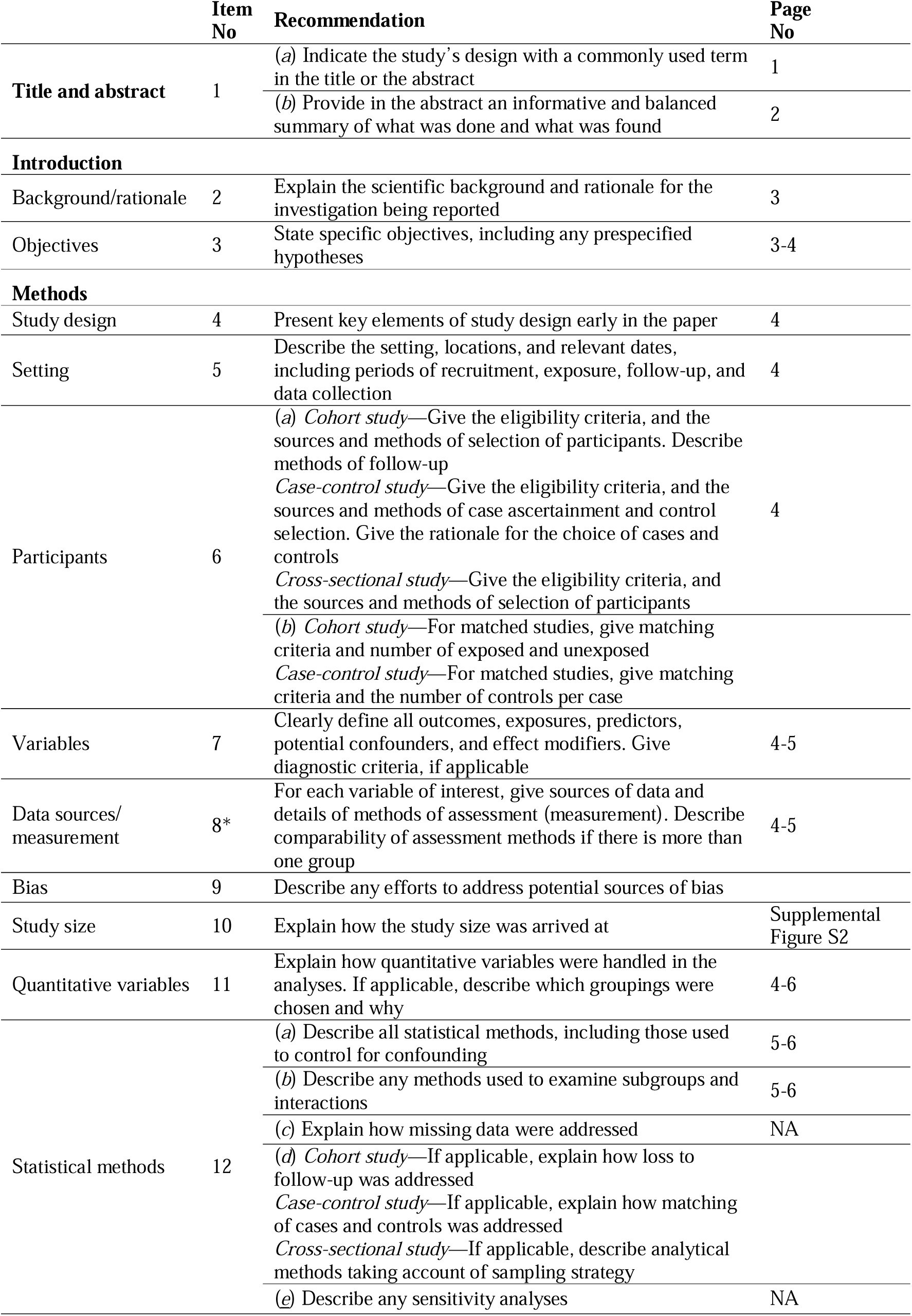
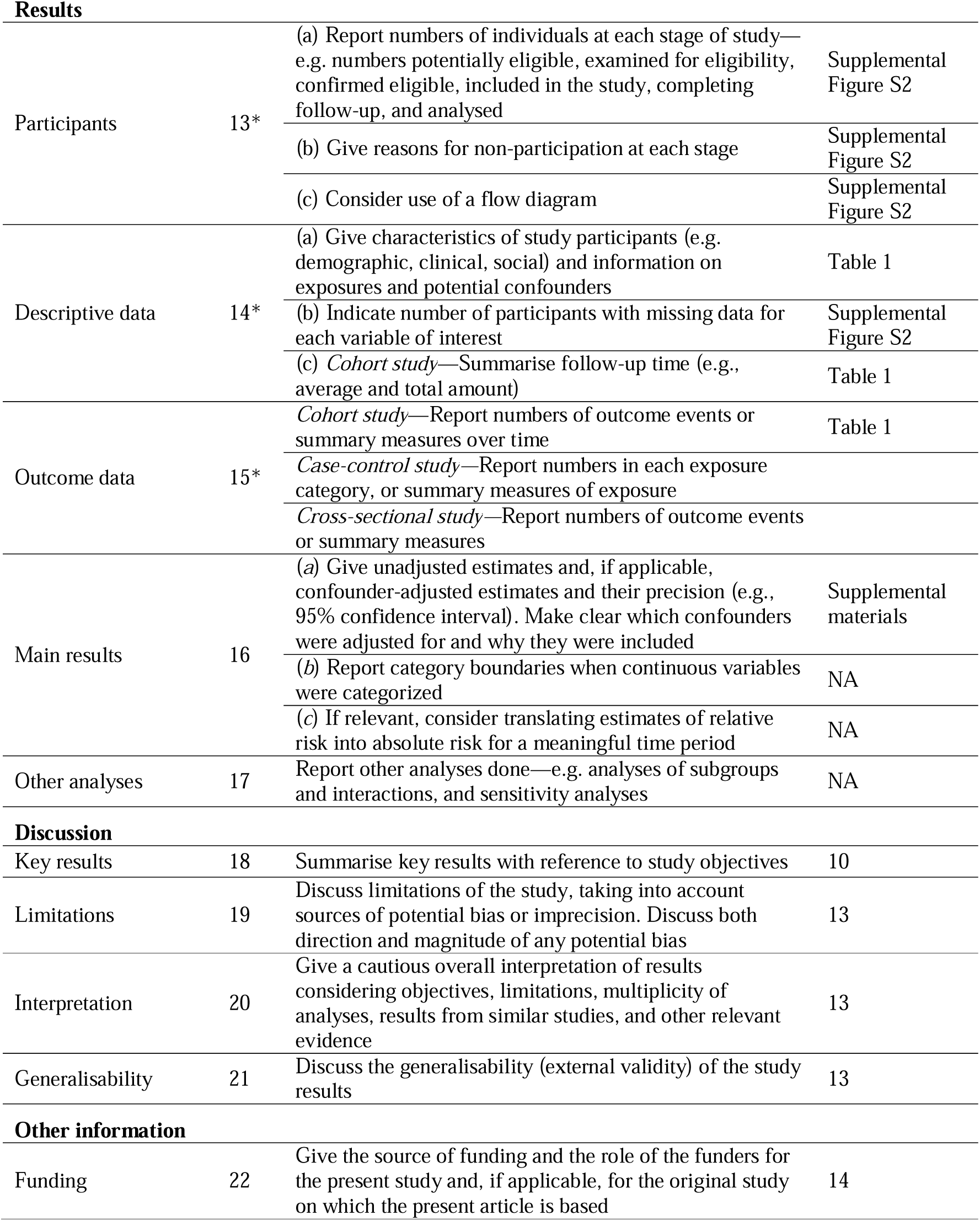
STROBE Statement—Checklist of items that should be included in reports of *cohort studies*

**Supplementary Table S2.** ICD-10 codes for Cancer.

| **Cancer Types and Subtypes** | **ICD-10 Code** |
| --- | --- |
| **Blood** | C81-C86, C88, C90-C96 |
| **Breast** | C50 |
| **Colorectal** | C18, C19, C21 |
| **Endocrine Gland** | C73-C75 |
| **Gastrointestinal** |  |
| Oesophagus | C15 |
| Stomach | C16 |
| Small Intestine | C17 |
| Liver & Intrahepatic Bile Ducts | C22 |
| Gallbladder & Biliary Duct | C23, C24 |
| Pancreas | C25 |
| Ill-defined Digestive Organs | C26 |
| **Genitourinary** |  |
| Kidney | C64 |
| Renal Pelvis & Ureter | C65, C66 |
| Bladder | C67 |
| Testis | C62 |
| Unspecified Urinary Organs | C68 |
| **Gynaecological** |  |
| Cervix | C53 |
| Ovary | C56 |
| Uterus | C54, C55 |
| **Head & Neck** |  |
| Lip, Oral, Cavity & Pharynx | C01-C14 |
| Sinuses, Larynx & Trachea | C31-C33 |
| **Lung** | C34 |
| **Melanoma** | C43 |
| **Skin (non-Melanoma)** | C44 |
| **Prostate** | C61 |
| **Other Cancer** |  |
| Bone & Articular Cartilage | C40, C41 |
| Central Nervous System | C69-C72 |
| Ill-defined Sites in Respiratory System | C30, C37-C39 |
| Mesothelioma | C45 |
| Other Neoplasms | C46-C49, C51, C52, C57, C58, C60, C63, C76, C80, C97 |
| **Multiple Cancer** | See *Notes*. |

**Notes.** For participants with more than one types of cancer, if they had multiple cancer subtypes (e.g., *Cervix* and *Ovary*) which fell under the same cancer type (e.g., *Gynaecological*) they were included in that cancer type (i.e., *Gynaecological*), not *Multiple Cancer*. If participants had multiple cancer subtypes (e.g., *Cervix* and *Kidney*) from different cancer types (e.g., *Gynaecological* and *Genitourinary*) they were placed in *Multiple Cancer*. Cancer subtypes which did not fall under a specific cancer type were included in *Other Cancer*.

**Supplementary Table S3.** ICD-10 Codes For Other Chronic Conditions.

| ICD-10 Chronic Disease Subgroups | ICD-10 Code |
| --- | --- |
| **Infectious and Parasitic Disease** |  |
| Tuberculosis | A15-A19 |
| Chronic viral hepatitis | B18 |
| Human immunodeficiency viruses (HIV) | B20-B23 |
| **Disease of the Circulatory System** |  |
| Chronic rheumatic heart disease | I05-I09 |
| Essential, secondary and all hypertension disease | I10-I13, I15 |
| Angina | I20 |
| Myocardial infarction | I21, I22 |
| Atherosclerotic heart disease | I25 |
| Pulmonary embolism | I26 |
| All valve disease | I34-I37 |
| Cardiomyopathy | I42 |
| All arrhythmia (excluding atrial fibrillation) | I44, I45, I47, I49 |
| Heart failure | I50 |
| Haemorrhagic stroke | I61, I62 |
| Thromboembolic stroke | I63 |
| Aneurysms | I71, I72 |
| Arterial thromboembolism | I74 |
| Venous thromboembolism | I81, I82 |
| **Mental and Behavioural Disorders** |  |
| Mental and behavioural disorders due to use of alcohol | F10 |
| Schizophrenia | F20 |
| Bipolar disorder | F31 |
| Depressive disorder | F32, F33 |
| Anxiety disorder | F40, F41 |
| Obsessive-compulsive disorder | F42 |
| Reaction to severe stress, and adjustment disorder | F43 |
| Eating disorders | F50 |
| Specific personality disorders | F60 |
| **Diseases of the Eye and Adnexa** |  |
| Cataracts | H25, H26 |
| Disorders of choroid and retina | H30, H31, H33-H36 |
| Glaucoma | H40, H42 |
| Disorders of ocular muscles and binocular movement | H49-H51 |
| Visual disturbances and blindness | H53, H54 |
| **Diseases of the Ear and Mastoid Processes** |  |
| Inner ear disorders | H80, H81, H83 |
| Hearing loss | H90, H91 |
| **Diseases of Blood and Blood-forming Organs and certain Disorders Involving the Immune mechanism** |  |
| Nutritional anaemias | H50-H53 |
| Haemolytic anaemias | B55-B59 |
| Aplastic anaemias | D60, D61 |
| Anaemia of chronic disease | D63 |
| Coagulation defects | D66-D68 |
| Platelet disorders | D69 |
| Immunodeficiency disorders (excluding HIV) | D70, D80-D84 |
| Sarcoid | D86 |
| **Diseases of the Digestive System** |  |
| Gastro-oesophageal reflux disorder | K21 |
| Other diseases of the oesophagus | K22 |
| Peptic ulcer disease | K25-K28 |
| Gastritis and duodenitis | K29 |
| Functional intestinal disorders | K30, K58, K89, K90 |
| Appendicitis | K35-K37 |
| Hernia | K40-K46 |
| Regional enteritis | K50 |
| Ulcerative colitis | K51 |
| Angiodysplasia of colon | K55 |
| Diverticular disease of intestine | K57 |
| Fatty liver disease | K76 |
| Chronic cholecystitis | K81 |
| Chronic pancreatitis | K86 |
| Coeliac disease | K90 |
| **Diseases of the Genitourinary System** |  |
| Chronic renal failure | N18, N19 |
| Urolithiasis | N20, N21 |
| Cystitis | N30 |
| Neuromuscular dysfunction of bladder | N31 |
| Hyperplasia of prostate | N40 |
| Chronic prostatitis | N41 |
| Hydrocele | N43 |
| Male infertility | N46 |
| Benign mammary dysplasia | N60 |
| Inflammatory disorders of breast | N61 |
| Disorders of breast and inflammation of female pelvic organs | N65, N66, N70, N73 |
| Chronic salpingitis and oophoritis | N70 |
| Inflammatory disease of uterus (excluding cervix) | N71 |
| Inflammatory disease of the cervix | N72 |
| Endometriosis | N80 |
| Female genitals prolapse | N81 |
| Fistulae involving female genital tract | N82 |
| Dysplasia of cervix uteri | N87 |
| Excessive, frequent and irregular menstruation | N92 |
| Female infertility | N97 |
| **Diseases of the Musculoskeletal System and Connective Tissues, Endocrine, Nutritional, and Metabolic Diseases** |  |
| Hypothyroidism | E01, E03 |
| Non-toxic goitre | E04 |
| Thyrotoxicosis | E05 |
| Diabetes mellitus | E10, E11, E13, E14 |
| Hyperparathyroidism | E21 |
| Acromegaly and gigantism | E22 |
| Hypopituitarism | E23 |
| Cushing syndrome | E24 |
| Adrenocortical insufficiency | E27 |
| Disorders of lipoprotein metabolism | E78 |
| Rheumatoid arthritis | M05, M06 |
| Crystal arthropathies | M10, M11 |
| Arthroses | M15-M19 |
| Meniscal and ligament damage of the knee | M53 |
| Dorsalgia & dorsopathies | M54 |
| Shoulder lesions | M75 |
| Osteoporosis | M80, M81 |
| **Diseases of the Central Nervous System (CNS)** |  |
| Chronic suppurative neurological disease | G06 |
| Systemic atrophies primarily affecting the CNS | G10-G14 |
| Extrapyramidal and movement disorders | G20, G21, G23, G25 |
| Other degenerative disease of CNS | G30, G31 |
| Multiple sclerosis | G35 |
| Other demyelinating diseases of CNS | G36, G37 |
| Epilepsy | G40 |
| Migraine | G43 |
| Transient ischaemic attack | G45 |
| Sleep disorders | G47 |
| Mononeuropathy, nerve root, plexus disorders of lower limbs | G54, G57 |
| Polyneuropathy | G60-G63 |
| Myoneural disorders | G70, G73 |
| Primary disorders of muscles | G71 |
| Paralytic syndromes affecting lower limbs | G80-G83 |
| Hydrocephalus | G91 |
| Spinal cord disease | G95 |
| **Diseases of the Respiratory System** |  |
| Allergic rhinitis | J30 |
| Chronic rhinitis, nasopharyngitis and pharyngitis | J31 |
| Chronic sinusitis | J32 |
| Chronic laryngitis | J37 |
| Chronic bronchitis | J41-J44 |
| Asthma | J45, J46 |
| Bronchiectasis | J47 |
| Occupational lung disease | J60-J68 |
| Interstitial pulmonary disease | J70, J84 |
| Chronic suppurative and necrotic of lower respiratory tract | J85, J86 |

**Supplementary Table S4.** UK Biobank variables used in the study.

| **Variable** | **UKB Biobank field** |
| --- | --- |
| Age at recruitment | p21022 |
| Sex | p31 |
| White ethnicity | p21000 |
| Body Mass Index | p21001 |
| Education/qualification attainment (any) | p6138 |
| Employment | p6142 |
| Townsend deprivation index | p22189 |
| Smoking | p20116 |
| Alcohol intake | p20117 |
| 24h movement behaviours |  |
| Sleep period | p40046 |
| Sedentary behaviour | p40047 |
| Light physical activity | p40048 |
| Moderate-to-vigorous physical activity | p40049 |
| Accelerometer |  |
| Start time of wear | p90010 |
| Data quality, good wear time | p90015 |
| Data problem indicator | p90002 |
| Cancer register |  |
| Date of cancer diagnosis | p40005 |
| Type of cancer: ICD10 | p40006 |
| First occurrence' of any code mapped to 3-character ICD-10 | p1712 |

**Supplementary Table S5.** 24h behaviour profiles (in raw minutes) in individuals with cancer (*N* = 10 152).

|  | **N (%)** | **MVPA**<br>*Mdn (25<sup>th</sup>, 75<sup>th</sup>)* | **LPA**<br>*Mdn (25<sup>th</sup>, 75<sup>th</sup>)* | **Sedentary**<br>*Mdn (25<sup>th</sup>, 75<sup>th</sup>)* | **Sleep period**<br>*Mdn (25<sup>th</sup>, 75<sup>th</sup>)* |
| --- | --- | --- | --- | --- | --- |
| **AT RECRUITMENT** |  |  |  |  |  |
| **Sex** |  |  |  |  |  |
| Female | 5 660 (55.8%) | 25 (11, 46) | 313 (252, 378) | 551 (482, 618) | 532 (491, 576) |
| Male | 4492 (44.2%) | 38 (19, 64) | 262 (206, 325) | 586 (514, 657) | 530 (486, 582) |
| **Ethnicity** |  |  |  |  |  |
| White | 10 016 (98.7%) | 30 (14, 54) | 290 (229, 357) | 565 (495, 635) | 531 (489, 579) |
| Non-white | 136 (1.3%) | 26 (11, 48) | 293 (239, 372) | 578 (484, 637) | 522 (472, 582) |
| **Body mass index** |  |  |  |  |  |
| Under to Normal weight (< 25 kg/m<sup>2</sup>) | 4 027 (38.8%) | 37 (19, 60) | 310 (248, 378) | 542 (473, 610) | 530 (491, 575) |
| Overweight (25.0 to 29.9 kg/ m<sup>2</sup>) | 4 355 (42.0%) | 31 (15, 54) | 284 (227, 348) | 572 (501, 640) | 532 (489, 579) |
| Obese (30+ kg/m<sup>2</sup>) | 1 987 (19.2%) | 19 (7, 39) | 261 (207, 330) | 605 (529, 675) | 531 (486, 586) |
| **Education/qualification attainment** |  |  |  |  |  |
| None | 1 079 (10.6%) | 25 (10, 45) | 290 (228, 360) | 556 (473, 626) | 546 (502, 604) |
| Any | 9 073 (89.4%) | 31 (14, 55) | 290 (230, 357) | 566 (497, 637) | 529 (488, 576) |
| **Employment** |  |  |  |  |  |
| None | 4 589 (45.2%) | 27 (12, 51) | 286 (227, 351) | 566 (496, 635) | 536 (495, 586) |
| Any | 5 563 (54.8%) | 33 (16, 56) | 293 (232, 363) | 564 (493, 636) | 527 (485, 573) |
| **Townsend deprivation index** |  |  |  |  |  |
| Least deprived (< -3.8) | 2 755 (27.1%) | 31 (15, 55) | 293 (233, 358) | 561 (493, 631) | 532 (490, 577) |
| Second least deprived (-3.8 to -2.5) | 2 557 (25.2%) | 31 (14, 54) | 296 (235, 362) | 561 (489, 632) | 531 (490, 578) |
| Second most deprived (-2.5 to -0.2) | 2 640 (26.0%) | 30 (13, 54) | 288 (229, 357) | 566 (494, 633) | 533 (492, 581) |
| Most deprived (> -0.2) | 2 200 (21.7%) | 30 (13, 53) | 283 (220, 351) | 575 (504, 646) | 527 (485, 580) |
| **Smoking** |  |  |  |  |  |
| Never | 5 516 (54.3%) | 32 (15, 55) | 295 (233, 362) | 560 (490, 632) | 531 (489, 578) |
| Previous | 40 30 (39.7%) | 29 (13, 54) | 286 (229, 354) | 569 (499, 637) | 531 (489, 579) |
| Current | 606 (6.0%) | 22 (9, 46) | 272 (206, 341) | 580 (503, 659) | 535 (494, 589) |
| **Alcohol consumption** |  |  |  |  |  |
| Never | 273 (2.7%) | 21 (9, 41) | 301 (243, 376) | 559 (483, 625) | 538 (498, 585) |
| Previous | 259 (2.6%) | 23 (10, 46) | 284 (207, 356) | 581 (500, 654) | 525 (487, 576) |
| Current | 9 620 (94.8%) | 31 (14, 55) | 290 (229, 357) | 565 (495, 635) | 531 (489, 578) |
| **AT THE START OF ACCELEROMETRY** |  |  |  |  |  |
| **Age** |  |  |  |  |  |
| <65 years old | 3542 (34.9%) | 34 (16, 57) | 293 (234, 367) | 561 (489, 635) | 526 (486, 572) |
| 65+ years old | 6610 (65.1) | 28 (13, 52) | 288 (227, 353) | 568 (497, 636) | 534 (491, 582) |
| **Time since cancer diagnosis** |  |  |  |  |  |
| < 1 year | 971 (9.6%) | 28 (12, 53) | 278 (217, 341) | 577 (503, 639) | 536 (493, 587) |
| 1 to 5 years | 3 451 (34.0%) | 31 (15, 56) | 285 (227, 354) | 567 (496, 637) | 531 (489, 579) |
| > 5 years | 5 730 (56.4%) | 30 (13, 53) | 295 (233, 362) | 562 (492, 634) | 530 (488, 577) |
| **Cancer type** |  |  |  |  |  |
| Blood | 412 (4.1%) | 27 (10, 45) | 266 (208, 350) | 580 (512, 652) | 533 (489, 586) |
| Breast | 1 911 (18.8%) | 25 (11, 47) | 310 (248, 382) | 554 (484, 624) | 530 (488, 572) |
| Colorectal | 391 (3.9%) | 26 (13, 45) | 296 (235, 352) | 568 (494, 638) | 532 (490, 583) |
| Endocrine Gland | 79 (0.8%) | 31 (13, 48) | 311 (250, 355) | 561 (511, 608) | 535 (497, 564) |
| Gastrointestinal Tract | 100 (1.0%) | 21 (10, 44) | 272 (200, 339) | 597 (519, 666) | 531 (468, 600) |
| Genitourinary | 296 (2.9%) | 31 (14, 57) | 266 (212, 340) | 582 (522, 659) | 526 (483, 577) |
| Gynaecological | 401 (3.9%) | 24 (9, 45) | 311 (250, 370) | 559 (491, 631) | 525 (490, 572) |
| Head and Neck | 115 (1.1%) | 35 (13, 54) | 281 (227, 362) | 569 (476, 650) | 529 (472, 576) |
| Lung | 57 (0.6%) | 15 (4, 37) | 288 (197, 345) | 580 (518, 674) | 538 (496, 587) |
| Melanoma | 395 (3.9%) | 33 (15, 56) | 292 (238, 354) | 559 (493, 629) | 529 (488, 574) |
| Multiple Primary | 1 714 (16.9%) | 27 (13, 52) | 284 (224, 351) | 569 (498, 636) | 534 (490, 588) |
| Other Cancer | 148 (1.5%) | 35 (15, 58) | 266 (213, 345) | 586 (510, 659) | 525 (484, 579) |
| Skin (non-melanoma) | 3 017 (29.7%) | 34 (17, 57) | 293 (232, 359) | 561 (491, 629) | 532 (492, 576) |
| Prostate | 1 116 (11.0%) | 41 (21, 65) | 266 (211, 328) | 579 (509, 653) | 531 (484, 581) |

*Notes.*

**Supplementary Table S6.** Unadjusted estimated minute differences in 24h behaviour profile across time since cancer diagnosis. Values are Bayesian posterior mean differences and 99% credible intervals. MVPA = moderate-to-vigorous physical activity, LPA = light physical activity. Bolded values indicate 99% credible intervals not containing 0.

|  | MVPA | LPA | Sedentary | Sleep Period |
| --- | --- | --- | --- | --- |
| **ref. Healthy** |  |  |  |  |
| **Cancer** | **-11.65**<br>[-12.72, -10.54] | **-13.19**<br>[-16.50, -9.74] | **10.40**<br>[6.46, 14.04] | **14.44**<br>[11.92, 17.00] |
| <1 year since diagnosis | **-12.70**<br>[-15.01, -10.38] | **-27.23**<br>[-35.51, -18.77] | **19.54**<br>[9.95, 29.44] | **20.39**<br>[13.87, 27.16] |
| 1-5 years since diagnosis | **-10.82**<br>[-12.29, -9.33] | **-16.54**<br>[-21.64, -11.74] | **11.99**<br>[6.55, 17.6] | **15.38**<br>[11.38, 19.16] |
| >5 year since diagnosis | **-11.97**<br>[-13.19, -10.72] | **-8.80**<br>[-12.82, -4.72] | **7.89**<br>[3.16, 12.29] | **12.87**<br>[9.83, 15.98] |
| **ref. Other Conditions** |  |  |  |  |
| **Cancer** | **-2.13**<br>[-2.86, -1.35] | **-6.61**<br>[-9.24, -3.72] | 1.52<br>[-1.73, 4.59] | **7.21**<br>[5.08, 9.40] |
| <1 year since diagnosis | **-3.18**<br>[-5.27, -0.96] | **-20.65**<br>[-28.59, -12.45] | **10.67**<br>[1.20, 19.97] | **13.16**<br>[6.58, 19.79] |
| 1-5 years since diagnosis | **-1.30**<br>[-2.49, -0.08] | **-9.96**<br>[-14.51, -5.48] | 3.11<br>[-1.79, 8.13] | **8.15**<br>[4.64, 11.54] |
| >5 year since diagnosis | **-2.45**<br>[-3.42, -1.49] | -2.22<br>[-5.95, 1.52] | -0.98<br>[-5.17, 3.07] | **5.65**<br>[2.99, 8.37] |
| **Pairwise across time since diagnosis** |  |  |  |  |
| 1-5 years since diagnosis<br>ref. <1 year since diagnosis | 1.88<br>[-0.78, 4.28] | **10.69**<br>[1.55, 19.87] | -7.55<br>[-17.91, 3.00] | -5.02<br>[-12.34, 2.04] |
| >5 year since diagnosis<br>ref. 1-5 years since diagnosis | -1.15<br>[-2.64, 0.31] | **7.74**<br>[2.16, 13.20] | -4.09<br>[-10.03, 1.93] | -2.50<br>[-6.62, 1.69] |
| >5 year since diagnosis<br>ref. <1 year since diagnosis | 0.73<br>[-1.64, 3.09] | 18.43<br>[9.27, 27.31] | -11.65<br>[-21.83, -1.00] | -7.52<br>[-14.51, -0.42] |

**Supplementary Table S7.** Unadjusted estimated minute differences in 24h behaviour profile across cancer types. Values are Bayesian posterior mean differences and 99% credible intervals. MVPA = moderate-to-vigorous physical activity, LPA = light physical activity. Bolded values indicate 99% credible intervals not containing 0.

|  | MVPA | LPA | Sedentary | Sleep Period |
| --- | --- | --- | --- | --- |
| **ref. Healthy** |  |  |  |  |
| Blood | **-15.46**<br>[-18.33, -12.26] | **-32.33**<br>[-44.41, -19.90] | **29.05**<br>[15.41, 43.74] | **18.75**<br>[9.05, 28.84] |
| Breast | **-16.06**<br>[-17.6, -14.46] | **7.64**<br>[1.02, 14.34] | -2.81<br>[-10.32, 4.35] | **11.23**<br>[6.26, 16.16] |
| Colorectal | **-14.61**<br>[-17.66, -11.04] | -8.67<br>[-21.32, 4.77] | 8.39<br>[-6.86, 23.19] | **14.89**<br>[4.68, 25.45] |
| Endocrine Gland | **-13.41**<br>[-20.05, -5.08] | 1.09<br>[-28.40, 34.28] | 6.00<br>[-26.60, 38.90] | 6.33<br>[-16.38, 29.30] |
| Gastrointestinal Tract | **-20.5**<br>2[-24.66, -15.21] | **-45.84**<br>[-68.93, -21.91] | **51.95**<br>[24.65, 81.01] | 14.41<br>[-5.70, 33.91] |
| Genitourinary | **-12.62**<br>[-16.36, -8.51] | **-33.66**<br>[-48.10, -18.83] | **33.37**<br>[15.97, 50.91] | **12.91**<br>[1.09, 25.19] |
| Gynaecological | **-18.06**<br>[-20.69, -15.13] | 1.90<br>[-11.99, 15.72] | 7.85<br>[-6.10, 22.89] | 8.31<br>[-1.58, 18.64] |
| Head and Neck | **-13.53**<br>[-18.90, -6.99] | -14.77<br>[-38.17, 9.05] | 17.08<br>[-10.80, 44.01] | 11.21<br>[-7.37, 29.95] |
| Lung | **-24.41**<br>[-28.21, -19.07] | **-44.56**<br>[-75.52, -10.79] | **41.39**<br>[4.10, 79.69] | **27.57**<br>[0.37, 56.08] |
| Melanoma | **-9.34**<br>[-13.05, -4.89] | -5.54<br>[-19.89, 8.48] | 2.40<br>[-12.01, 17.26] | **12.47**<br>[2.91, 22.00] |
| Multiple Primary | **-13.24**<br>[-14.99, -11.51] | **-20.05**<br>[-26.6, -13.41] | **14.66**<br>[7.20, 22.17] | **18.64**<br>[13.47, 24.07] |
| Other Cancer | **-11.85**<br>[-17.06, -5.41] | **-27.14**<br>[-46.39, -6.31] | **32.36**<br>[10.28, 55.72] | 6.63<br>[-9.35, 22.62] |
| Skin (non-melanoma) | **-7.53**<br>[-9.18, -5.73] | **-11.08**<br>[-16.36, -5.83] | 3.81<br>[-2.09, 9.77] | **14.80**<br>[10.81, 18.78] |
| Prostate | -2.61<br>[-5.52, 0.50] | **-35.03**<br>[-42.73, -27.26] | 25.29<br>[16.02, 33.98] | **12.36**<br>[6.19, 18.84] |
| **ref. Other Conditions** |  |  |  |  |
| Blood | **-5.95**<br>[-8.61, -2.94] | **-25.71**<br>[-37.63, -13.18] | **20.13**<br>[6.35, 34.64] | **11.53**<br>[1.83, 21.77] |
| Breast | **-6.55**<br>[-7.87, -5.17] | **14.27**<br>[8.05, 20.82] | **-11.73**<br>[-18.81, -4.81] | 4.02<br>[-0.76, 8.88] |
| Colorectal | **-5.10**<br>[-8.21, -1.63] | -2.05<br>[-14.81, 11.54] | -0.52<br>[-15.49, 14.39] | 7.67<br>[-2.54, 17.98] |
| Endocrine Gland | -3.90<br>[-10.45, 4.49] | 7.71<br>[-21.53, 41.08] | -2.92<br>[-36.06, 29.67] | -0.89<br>[-23.62, 22.1] |
| Gastrointestinal Tract | **-11.01**<br>[-15.06, -5.79] | **-39.22**<br>[-62.16, -15.21] | **43.03**<br>[15.88, 72.51] | 7.19<br>[-12.51, 26.45] |
| Genitourinary | -3.11<br>[-6.73, 0.85] | **-27.04**<br>[-41.53, -12.76] | **24.45**<br>[7.41, 41.62] | 5.70<br>[-5.94, 17.71] |
| Gynaecological | **-8.55**<br>[-11.02, -5.77] | **8.53**<br>[-5.48, 22.56] | **-1.07**<br>[-15.1, 13.92] | 1.09<br>[-8.84, 11.17] |
| Head and Neck | -4.01<br>[-9.28, 2.45] | -8.14<br>[-31.28, 15.79] | 8.16<br>[-19.55, 34.86] | 3.99<br>[-14.76, 22.68] |
| Lung | **-14.89**<br>[-18.58, -9.61] | **-37.93**<br>[-68.54, -4.46] | 32.47<br>[-4.76, 71.08] | 20.35<br>[-6.75, 48.70] |
| Melanoma | 0.17<br>[-3.43, 4.67] | 1.09<br>[-12.89, 14.79] | -6.52<br>[-20.76, 8.20] | 5.26<br>[-4.27, 14.72] |
| Multiple Primary | **-3.72**<br>[-5.32, -2.12] | **-13.43**<br>[-19.43, -7.18] | 5.74<br>[-1.48, 12.74] | **11.42**<br>[6.64, 16.57] |
| Other Cancer | -2.34<br>[-7.57, 3.97] | -20.51<br>[-39.84, 0.36] | **23.44**<br>[1.37, 46.91] | -0.59<br>[-16.32, 15.43] |
| Skin (non-melanoma) | **1.98**<br>[0.51, 3.56] | -4.45<br>[-9.39, 0.41] | -5.11<br>[-10.61, 0.35] | **7.58**<br>[3.80, 11.42] |
| Prostate | **6.90**<br>[4.07, 9.95] | **-28.41**<br>[-35.79, -20.95] | **16.37**<br>[7.36, 24.91] | 5.14<br>[-1.17, 11.14] |

**Supplementary Figure S1.**
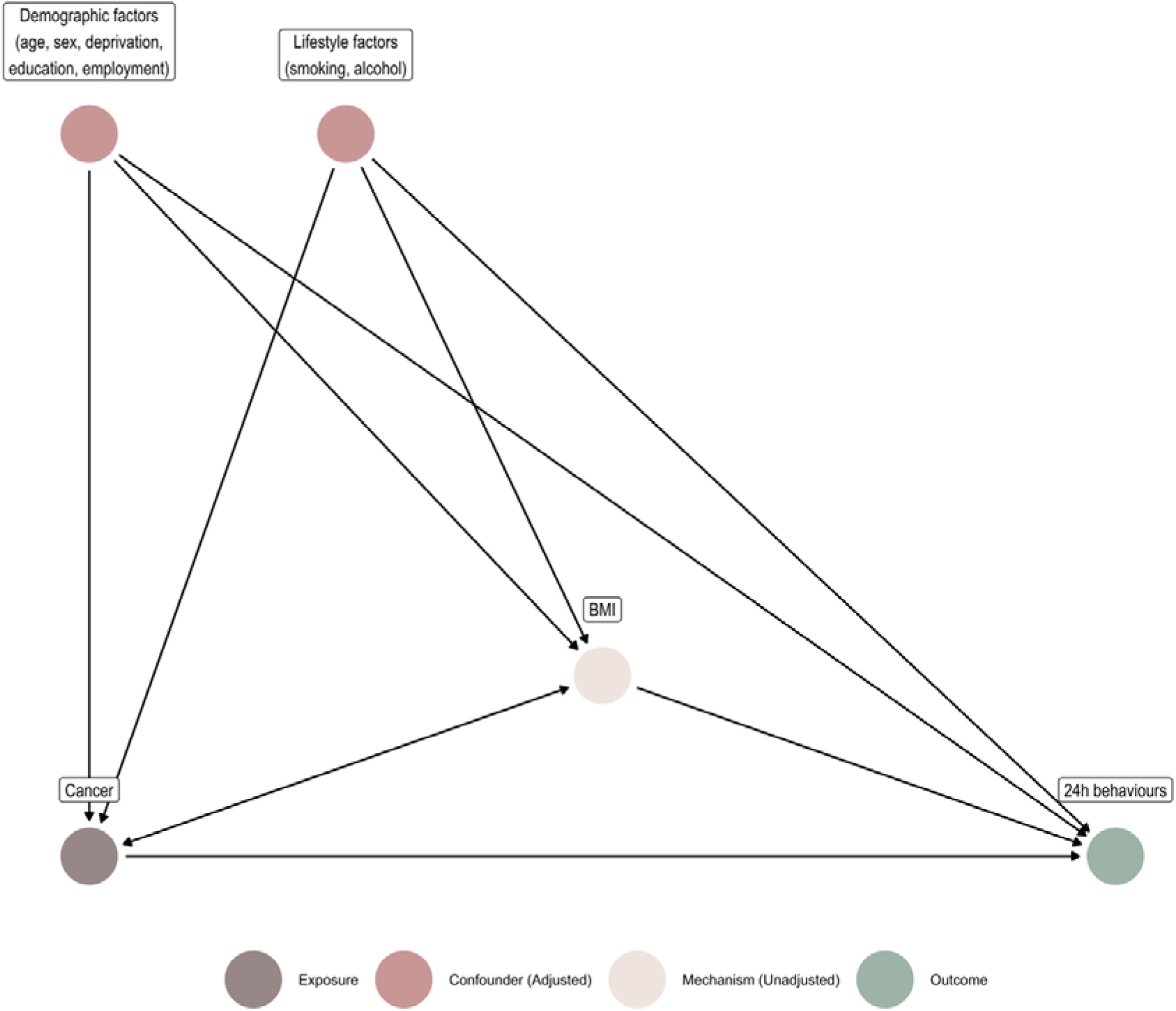
Directed acyclic graph.

**Supplementary Figure S2.**
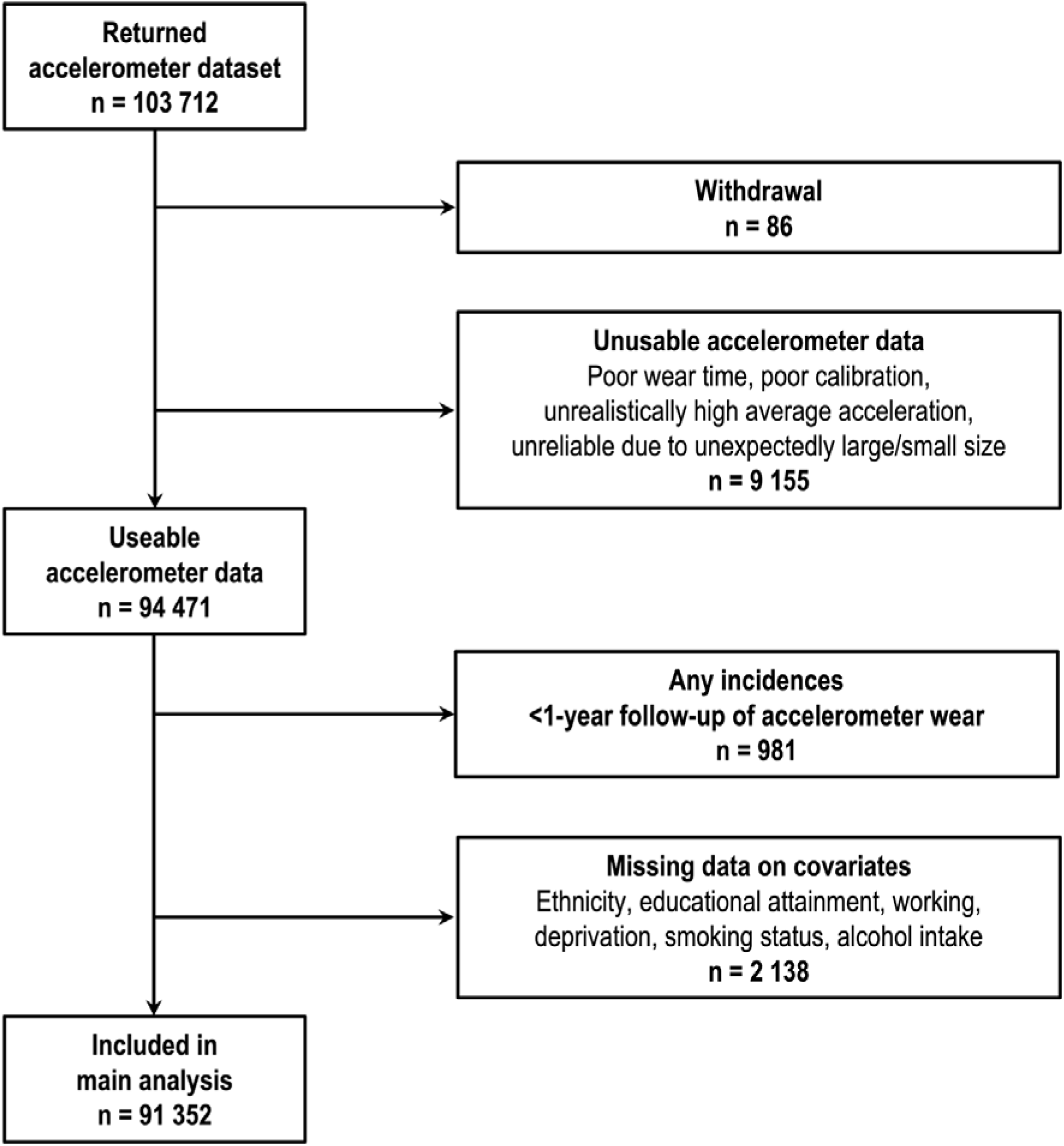
Participant flow diagram for data analysis.

**Supplementary Figure S3.**
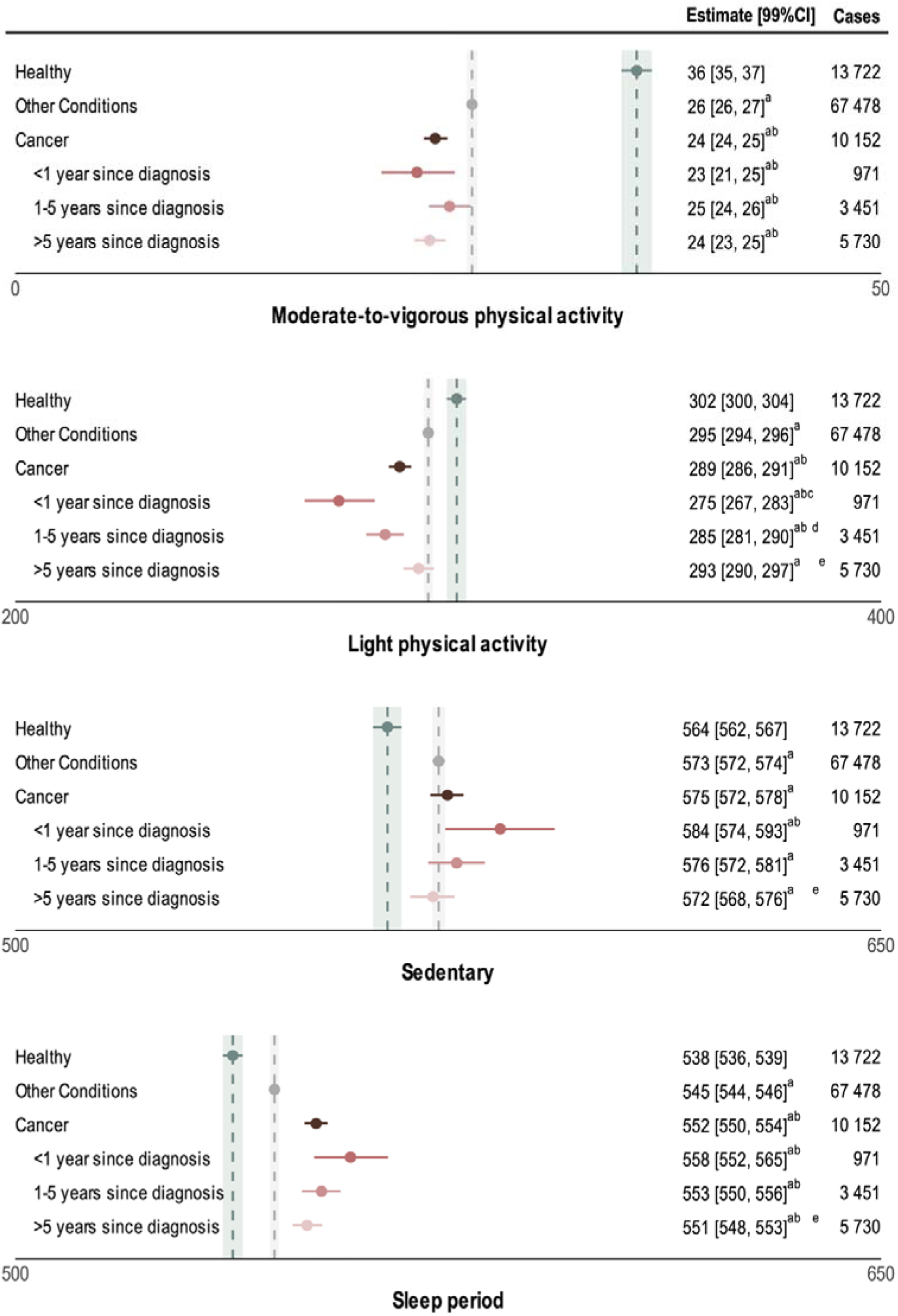
Unadjusted estimated 24h behaviour profile (in minutes) across time since cancer diagnosis. Values are Bayesian posterior mean differences and 99% credible intervals. Vertical dashed lines represent estimated means, and shaded regions are 99% credible intervals, of non-cancer groups (green = healthy, grey = other conditions). ^a^significant difference from *Healthy*, ^b^significant difference from *Other Conditions*, ^c^significant difference between <1 year and 1-5 years since diagnosis, ^d^significant difference between 1-5 and >5 years since diagnosis, ^e^significant difference between <1 year and >5 years since diagnosis.

**Supplementary Figure S4.**
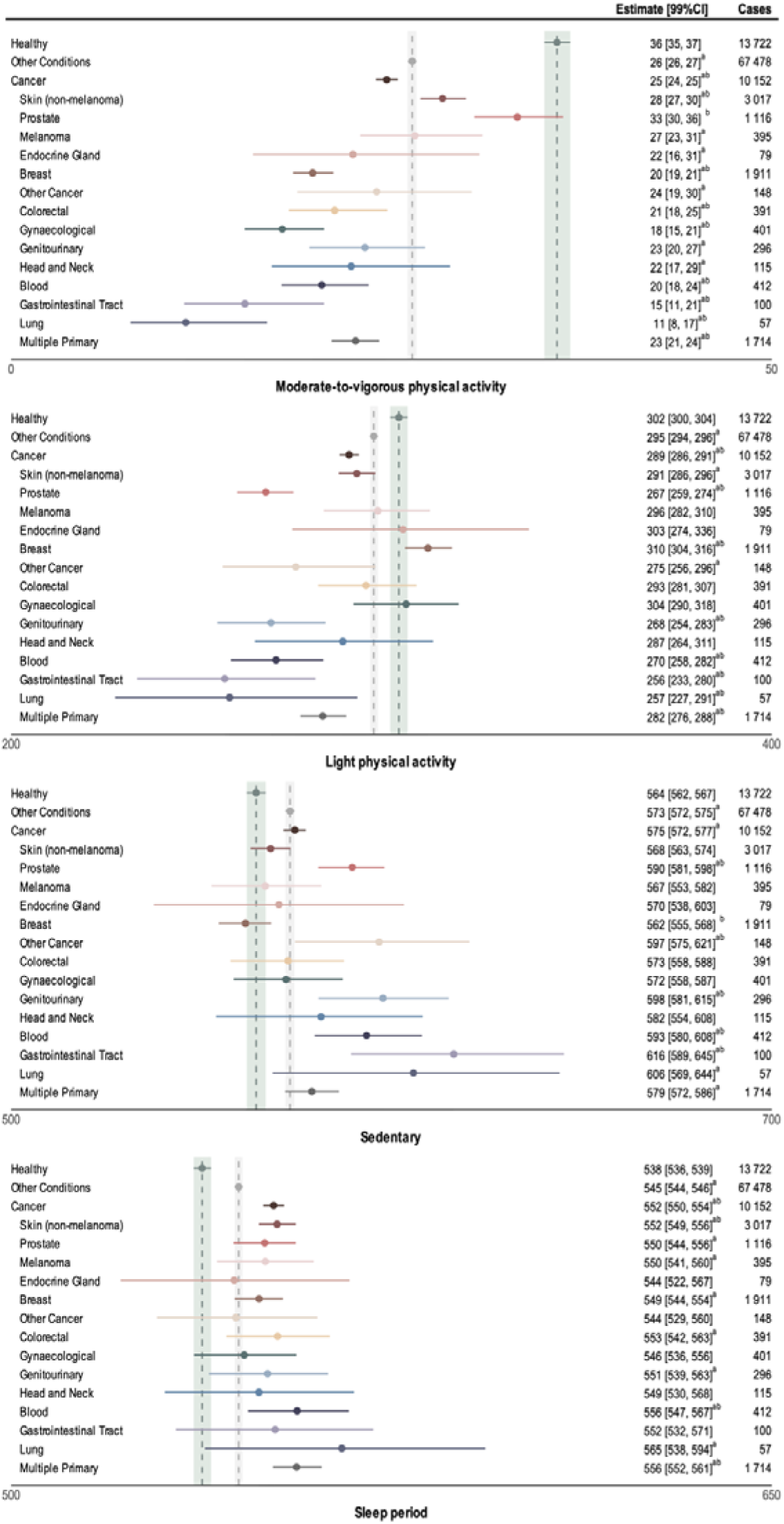
Unadjusted estimated 24h behaviour profile (in minutes) across cancer types. Values are Bayesian posterior mean differences and 99% credible intervals. Vertical dashed lines represent estimated means, and shaded regions are 99% credible intervals, of non-cancer groups (green = healthy, grey = other conditions). ^a^significant difference from *Healthy*, ^b^significant difference from *Other Conditions*.

